# Observational study of HPV genitalia and oral infection in an unvaccinated population of men who have sex with men infected with HIV in Northwest Spain

**DOI:** 10.1101/2021.08.09.21261786

**Authors:** Alexandre Pérez-González, Sonia Pérez, Raquel Carballo, Elena López-Diez, Jacobo Limeres-Posse, Antonio Ocampo

**Author notes:** Corresponding author: Alexandre Pérez González, Galicia Sur Health Research Institute, Virology and Pathogenesis Group. Internal Medicine Department. Álvaro Cunqueiro Hospital, St Clara Campoamor N° 341. Vigo 36312, Spain.

## Abstract

**Background:** Human papillomavirus (HPV) infection is the most common sexual transmitted disease (STD) and a risk factor for penile, oral and anal cancer. Human immunodeficiency virus (HIV) coinfection increases the risk of cancer. While HPV anal infection is well studied in men-who-have-sex-with-men (MSM), HPV genitalia and oral infection is less known.

**Methods:** This prevalence study of HPV infection in genitalia and oral cavity in HIV-MSM patients included 107 HPV non-vaccinated subjects. HPV-DNA was detected with Anyplex™ II HPV28 method. Participants completed a questionnaire on lifestyle and sexual behavior.

**Results:** Median age was 43 years (range 35-54 years); 97 patients were on antiretroviral treatment (ART); 81 (75.7%) had undetectable HIV-RNA; median CD4-lymphocyte count was 746 cell/mm^3^; 70 (65.4%) participants had a previous STD. HPV was detected in genitalia in 37 (34.6%) subjects; 26 (24.3%) in oral cavity and 12 (11.2%) in both locations. High-risk HPV genotypes were detected in 24 (22.4%) patients in genitalia and 15 (14%) in oral cavity.

**Conclusions:** HPV infection is common in unvaccinated HIV-MSM patients. Detectable HIV-RNA was associated with higher HPV prevalence in genitalia. High oncogenic risk HPV genotypes were more common in genitalia than in mouth.

**Summary text:** HPV infection is common in HIV infected subjects and it is a risk factor for many types of cancer. Although anal conduct is the most studied location, HPV can also infect genitalia and oral cavity. However, the frequency and distribution of HPV strains is different in both locations.

## Introduction

Human papillomavirus (HPV) infection is the most common sexual transmitted disease (STD) in many countries, including United States [1] and some South European countries [2]. HPV is the main cause of cervical cancer in woman [3] and it is associated with an increased risk of penile [4], anal [5] and oral cancer [6]. HPV strains are classified based on their oncogenic risk [7]. While some genotypes are associated with genital warts (genotypes 6 and 11), others are closely related with dysplasia and cancer (such as HPV-16, HPV-18, HPV-31). Finally, some genotypes are probably associated with an increased risk of cancer (probable high risk), while others have an indeterminate relationship with dysplasia [7]. Human immunodeficiency virus (HIV) and HPV coinfection is frequent [8]. Both viruses share sexual intercourse as the main route of transmission [8]. Receptive anal intercourse is the sexual practice with higher STD transmission probability [8]. Despite the high efficacy of antiretroviral treatment (ART), the risk of anal cancer is increased in men who have sex with men (MSM) infected by HIV (HIV-MSM) [9]. In the recent years, anal dysplasia screening programs have been developed due to a growing anal cancer incidence [10]. HPV-related oropharyngeal cancer incidence is also rising up [11]. Finally, almost 50% of penile cancers are related to a HPV infection, being HPV-16 the most predominant [12].

Coinfection of HPV, HIV and other STDs is also frequent [13-14]. *Chlamydia trachomatis* coinfection may also increase HPV-related cancer in males [15] and infertility [16]. However, it is unknown the interaction between HPV and other STD as well as its role on carcinogenesis and clinical relevance.

Nevertheless, prevalence, relevance and genotype distribution of HPV in MSM-HIV patients outside anal conduct remains unclear. Our study group found a prevalence of HPV oral infection 13.4% in HIV-MSM [30], but genitalia HPV infection rate remains unknown,

The aim of this study is to analyze the prevalence of HPV infection and genotype distribution outside anal conduct in an unvaccinated cohort of Northwest Spain.

## Methods

This is a prevalence study in HIV-MSM patients in the follow-up HIV unit at the Álvaro Cunqueiro Hospital, a third level hospital in Vigo, Northwest Spain (population area around 540,000 inhabitants). Subjects were recruited between January and December 2019. HPV-DNA was studied in genitalia and oral samples. A total of 107 HIV-MSM subjects were recruited. HPV-vaccinated subjects were excluded. Oral samples were taken after one hour fast. Patients were asked to make two rinses with 5 mL of sodium chloride. First rinse was safely disposed while the second was sent to laboratory in a sterile bottle at ambient temperature. Genitalia samples were obtained with a cytologic brush (Endobrush®, Covaca S.A., Madrid, Spain) from glans, coronal sulcus and scrotum, and stored in TrisEDTA pH8, molecular grade. Anal, oral and genitalia swabs were obtained for bacterial STD at the same time as HPV sampling and stored in PCR media (Roche Diagnostics®, Basel, Switzerland). We used two swabs per location, one for bacterial culture and another for *Chlamydia trachomatis* and *Neisseria gonorrhoeae* detection. Serum was obtained for *Treponema pallidum* serological study. Demographical and clinical data were obtained from medical records. Patients completed a questionnaire on lifestyle and sexual behavior.

HPV-DNA was extracted from genitalia and oral samples employing the QIAcube automated extraction system (Qiagen, Hilden, Germany). The HPV genotypes were identified by applying the Anyplex™ II HPV28 detection method (Seegene, Seoul, South Korea), following the manufacturer’s recommendations. This test simultaneously detects 19 types of high-risk HPV and 9 types of low-risk HPV. The system performs 3 real-time multiplex polymerase chain reactions per sample using DPO™ Technology and the TOCE™ technology melting curve analysis method.

Quantitative variables are expressed as median and interquartile range. Qualitative variables are shown as absolute value and percentages. Categorical variables were compared with χ-square test or Fischer exact test as needed. For quantitative variables comparison we used U Man Whitney test. A p value less than 0.05 was considered significant. Statistical analysis was performed with Statistical Package for Social Sciences (SPSS), IBM, version 22.

### Ethics

All patients signed an informed consent form. Study was approved by Ethics Committee of Pontevedra-Vigo-Ourense (reference 217/2019).

## Results

Clinical and epidemiological characteristics of the study group are shown in table 1. Most of them were Spanish (n=80, 74.8%). Median age was 43 years (IQR, 35-54); 97 of them (89.7%) were on ART while 10 were naïve at the time of recruitment. The most common ART regimen consisted in an integrase inhibitor (II) plus two nucleoside reverse transcriptase inhibitors (n=56, 59.7%). HIV classification was stage C for 21 participants (19.6%). In 25 patients (23.4%), the lowest CD4 lymphocyte count was under 200 cell/mm^3^. At the time of the recruitment, 81 subjects (75.5%) had an HIV RNA count under 50 copies/mL. Median time since HIV diagnosis to recruitment was 8 years (range 4—13); 64 subjects (59.8%) had a prior diagnosis of anal HPV infection (HPV-16 n=22; HPV-18 n=10). Most of the patients (n=70, 65.4%) had a previous STD, being syphilis the most common.

**Table 1.** Baseline characteristics of study population

|  |  |
| --- | --- |
| **Ethnicity, n (%)** |  |
| Spanish | 80 (74.8%) |
| Latin-American | 25 (23.4%) |
| Other | 2 (1.8%) |
| Age in years, median (range) | 43 (35 – 54) |
| Under HIV treatment | 97 (90.7%) |
| Naive to ART | 10 (9.3%) |
| **ART treatment** |  |
| Integrase inhibitor plus two nucleoside reverse transcriptase inhibitors | 56 (59.7%) |
| Non-nucleoside reverse transcriptase inhibitor plus two nucleoside reverse transcriptase inhibitors | 20 (21.3%) |
| Protease inhibitor plus two nucleoside reverse transcriptase inhibitors | 7 (7.4%) |
| Protease inhibitor plus lamivudine | 6 (6.4%) |
| Integrase inhibitor plus lamivudine | 2 (2.1%) |
| Protease inhibitor plus integrase inhibitor | 2 (2.1%) |
| Integrase inhibitor plus one non-nucleoside reverse transcriptase inhibitor | 1 (1.1%) |
| Nadir CD4 lymphocyte (cell/mm<sup>3</sup>; median and interquartile range) | 326 (197 – 456) |
| CD4 nadir less than 200 (cell/mm<sup>3</sup>), n (%) | 25 (23.4%) |
| CDC C stage, n (%) | 21 (19.6%) |
| CD4 lymphocyte at recruitment (cell/mm<sup>3</sup>; median and interquartile range) | 746 (447 – 966) |
| HIV RNA < 50 copies/mL | 81 (75.7%) |
| Time since HIV diagnosis (years; median and interquartile range) | 8 (4 – 13) |
| Circumcision, n (%) | 22 (20.6%) |
| Anogenital condylomata acuminata | 17 (15.9%) |
| Prior anal high grade intraepithelial dysplasia | 13 (12.1%) |
| Prior HPV anal infection, high risk genotype | 68 (63.6%) |
| **Tobacco use** |  |
| Active smoker, n (%) | 34 (31.8%) |
| Former smoker, n (%) | 19 (17.8%) |
| Never smoker, n (%) | 53 (49.5%) |
| Previous sexual transmitted disease, n (%) | 70 (65.4%) |
| Syphilis, n (%) | 64 (59.8%) |
| Gonorrhea, n (%) | 21 (19.6%) |
| *Chlamydia trachomatis* infection, n (%) | 10 (9.3%) |
| **Results of sexual behavior questionnaire** |  |
| Age of first sexual intercourse, median (interquartile range) | 17 (15 – 18) |
| Total number of sexual partners in life, median (interquartile range) | 100 (40 - 300) |
| Last year total number of sexual partners, median (interquartile range) | 4 (1 - 10) |
| Consistent condom use (penetration), n (%) | 55 (51.4%) |
| Consistent condom use (oral sex), n (%) | 5 (4.7%) |

### Sexual behavior questionnaire

Sexual behavior questionnaire results are summarized in table 1. Consistent use of condom in penetration was reported by 55 patients (51.4%), while on oral sex, the proportion dropped to 4.7% (n=5). Median number of total sexual partners lifetime was 100 (IQR,40-300) and 4 in the past year (IQR, 1-10). Median age of first sexual intercourse was 17 years (IQR, 15-18). The sexual behavior questionnaire is attached as supplementary material.

### HPV prevalence and distribution

HPV prevalence results are shown in figure 1. HPV DNA was detected in 37 genitalia samples (34.6%). Genotype distribution was as follows: high risk genotypes were detected in 24 subjects (22.4%), probable high risk in 12 patients (11.2%) and low risk in 21 (19.6%). HPV DNA was found in oral samples of 26 participants (24.3%). High risk genotypes were identified in 15 (14%) subjects, while probable high-risk strains were found in 9 (8.4%), low risk in 9 (8.4%) and indeterminate risk in 2 (1.9%).

**Figure 1.**
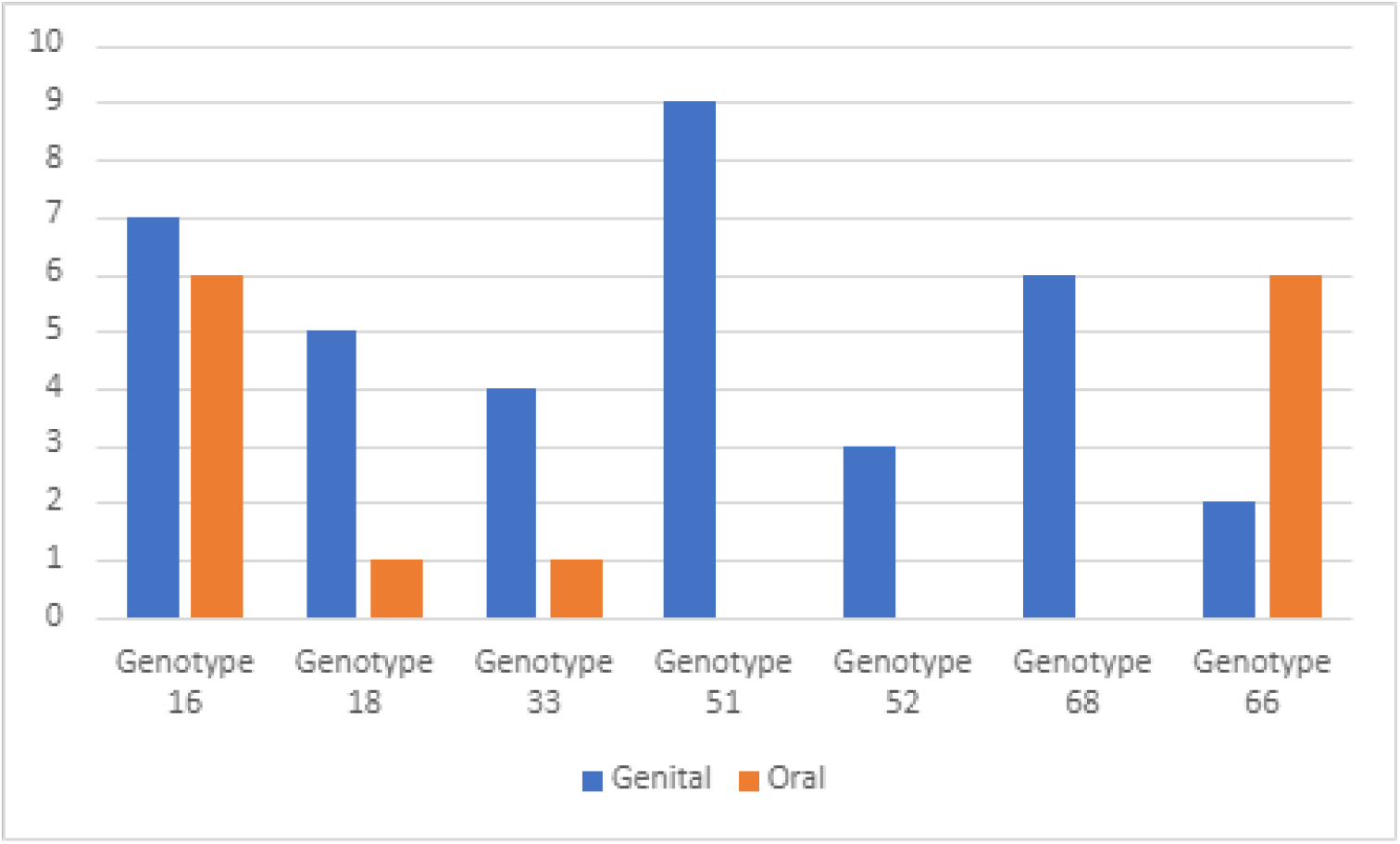
Frecuencies of most common HPV genotypes

Genotype prevalence was different between genitalia and oral samples. HPV-51 was the most common in genitalia samples (n=9; 8.4%), followed by HPV-16 (n=7, 6.5%) and HPV-68 (n=6, 5.6%). However, HPV-66 was the most frequent strain in oral samples, reaching 6 cases (5.6%) tied with HPV-16 (6 cases, 5.6%). HPV-51 and HPV-68 were not detected in oral samples. Coinfection of more than one HPV genotype was more common in genitalia than oral cavity (52.8% vs 30.8% respectively).

Prevalence of HPV genitalia infection was higher in patients with detectable HIV-RNA (65.4% vs 24.7%; p<0.001), but no difference was observed in HPV oral infection between suppressed and non-suppressed viral load (24.9% and 23.1%, respectively).

HPV genitalia infection did not vary between smokers and non-smokers. Regarding oral cavity, there was no significant differences in HPV prevalence in smokers and ex-smokers vs. never smokers (30.2% and 20.5%, respectively). HPV oral infection prevalence did not differ between patients with a history of a previous STD. In both locations, prevalence was not related with HIV stage C nor a CD4 cell count less than 200 cell/mm^3^. Age, number of lifetime sexual partners, consistent condom use, age of first sexual intercourse, CD4 lymphocyte count, nadir CD4 lymphocyte count and tobacco consumption did not differ between HPV positive and HPV negative subjects.

### Bacterial-STD samples

Bacterial study resulted positive in 21 (19.6%) subjects. The most frequent STD excluding HPV was *Chlamydia trachomatis* infection with 12 cases (11.2%), followed by *Neisseria gonorrhoeae* (9 cases, 8.4%) and *Haemophilus parainfluenzae* (2 subjects, 1.9%). *Chlamydia* spp. and gonococcus spp. coinfection was detected in 2 subjects (1.9%). Ten subjects (9.3%) tested positive for anal infection, followed by oral cavity (5 cases, 4.7%) and urethra (4 cases, 3.7%). One subject (0.9%) tested positive in two locations: anal conduct and oral cavity. Ten patients (9.3%) tested positive for syphilis. HPV prevalence did not vary between patients with a positive or a negative result for another STD (47.6% vs 46.5% respectively).

## Discussion

HPV is a major concern in HIV-MSM patients due to the increased risk of anal cancer. However, there is less knowledge about the frequency of HPV infection in genitalia and oral cavity.

HPV genital and oral prevalence were similar to those found in another European studies of HIV infected males [17-20] usually high, between 20% and 31%.

Overall prevalence of HPV infection was higher in genitalia than oral cavity. HPV prevalence in genitalia was also higher at each oncogenic risk category, except for 2 cases of indeterminate oncogenic risk of HPV in oral cavity [21]. A prior Dutch study showed a higher HPV prevalence in oral samples than in genitalia. We could not find any studies in Spanish population that compared HPV infection prevalence between oral and genitalia samples.

Distribution of genotypes also varied between genitalia and oral samples. Interactions between HPV and mucosal surface, microbiome and local immunity may be different in genitalia and in oral tissues. These interactions are yet poorly understood, but may play a role on infectivity and potential carcinogenesis of each genotype [22]

HPV-66 was the most common in oral samples. This genotype is classified as probable high risk oncogenic strain with an estimated prevalence in woman around 16% [23]. The prevalence and relevance of this genotype in HIV-MSM is still unknown.

HPV-16 was one of the most common genotypes in both locations. King et al. performed a meta-analysis, finding a HPV-16 prevalence around 3% in oral samples. The prevalence in our study is similar to previous research [24-25], slightly above the meta-analysis published by King [18]. HPV-16 is a major concern due to its role in head and neck [26], penile [27] and anal [28] cancer.

HPV-51 was the most common strain isolated from genital samples; however, it was not detected in any oral sample. HPV-51 is a high oncogenic risk genotype, which anal prevalence in a nationwide Spanish cohort was around 20% [29].

HPV-18, HPV-33, HPV-52 and HPV-68 were also more common in genitalia than oral samples. All of them are considered high oncogenic risk genotypes. Prevalence of each type is unknown in the Spanish population. Most of the previous studies have been focused on anal samples [29], while real prevalence and incidence of some genotypes outside anal conduct remains unknown.

Coinfection of multiple genotypes was also more common in genitalia than in oral cavity. HPV coinfection is frequent in HIV-MSM [30]. HIV enhance individual susceptibility to HPV infection through various mechanisms. Firstly, HIV decrease the number of lymphocytes CD4 count, which increases the risk of HPV infection. Secondly, cells exposed to HIV produce many inflammatory substances, damaging epithelial barriers and other defensive mechanisms [31].

We found no correlation between HPV genotypes in genitalia and in oral cavity. This also happened on previous studies [32]. Our previous research showed no concordance between anal and oral HPV genotype [33]. Therefore, we decided to only consider genotypes 16 and 18 in anal samples. Again, no concordance was found between anal and non-anal HPV genotypes.

Consistent use of condom was similar to a previous study of Tao et al in Chinese MSM-HIV population [34]. We did not find a relationship between condom use and HPV infection rate. Consistent use of condom during penetration was not associated with a lower HPV infection prevalence. This could be attributed to two factors. Firstly, HPV can be transmitted by direct contact with unprotected areas, such as scrotum, perineum and lips [35]. Secondly, low sample size may limit the statistical analysis. Use of condom during oral sex was very low. We did not find any study regarding the use of condom in oral sex in HIV-MSM. However, Hollway et al. found a rate use around 9% in males [36].

In our study, age, number of sexual partners, age of initial sexual intercourse and CD4 lymphocyte count were not associated with an increased HPV prevalence. A detectable HIV-RNA was associated with a higher prevalence on genitalia infection, but not within the in the oral cavity. Previous studies showed an increased risk of HPV anal and genitalia infection in patients with detectable HIV-RNA or low CD4 lymphocyte count [37-38].

## Conclusions

Genital and oral HPV infection is frequent in HIV-MSM, including high risk oncogenic HPV strains. HPV 16 was the most common genotype in the study population. Prevalence and distribution of genotypes varied based on anatomic location, but no correlation was found between genitalia and oral samples. A detectable HIV-RNA was associated with a higher HPV genital prevalence but not with HPV oral infection rate. CD4 lymphocyte count, age and number of sexual partners were not associated with a higher prevalence of HPV infection.

Our study has several limitations. Firstly, the study of HPV infection prevalence outside anal conduct is challenging due to lack of commercial validated kits and a clear standard of sample collection. Moreover, management of this specimens in the Microbiology Department does not follow well established indications. Each laboratory uses a customized routine in order to detect HPV-DNA. We used the most common technique for each location (triple sample collection in genitalia and rinse in oral cavity) but both procedures are not validated. These two elements could explain the great variability of results between different research studies. Our research group has used these two techniques in previous studies, increasing our experience in HPV-DNA detection outside anal conduct [32, 39-40]. Secondly, sample size is relatively small, so many comparisons could not be accurate (e.g., condom usage). Finally, only 10 patients were naive to ART, so comparisons between naive and treated subjects may not be significant due to low sample size.

## Supporting information

Questionnaire

## Data Availability

The data that support this study cannot be publicly shared due to ethical or privacy reasons and may be shared upon reasonable request to the corresponding author if appropriate.

## Acknowledgements

We would like to acknowledge Pedro Diz Dios (University of Santiago de Compostela) for writing and content review. Also, we would like to acknowledge Laura Piñeiro Lourés (Galicia Sur Health Research Institute) for providing language support.

