## Supplementary material for "Observational study of HPV genitalia and oral infection in an unvaccinated population of men who have sex with men infected with HIV in Northwest Spain": Questionnaire

**Supplementary material**
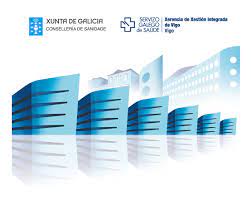
 **1**

Sexual behaviour questionnaire (*translated from original in Spanish*)

**HPV study 217/2019**

Subject ID:

Date of visit:

Sexual role:	Active	Pasive	Versatile

Age of first sexual intercourse:	(years)

Number of lifetime sexual partners:

Number of sexual partners last year:

Do you currently have a stable sexual partner?	Yes	No

Do you usually use condom during sexual intercourse?	Yes	No

Do you usually use condom during oral sex?	Yes	No

Are you circumcised?	Yes	No
